# Using Real-World Data to Rationalize Clinical Trials Eligibility Criteria Design: A Case Study of Alzheimer’s Disease Trials

**DOI:** 10.1101/2020.08.02.20166629

**Authors:** Qian Li, Yi Guo, Zhe He, Hansi Zhang, Thomas J George, Jiang Bian

## Abstract

Low trial generalizability is a concern. The Food and Drug Administration had guidance on broadening trial eligibility criteria to enroll underrepresented populations. However, investigators are hesitant to do so because of concerns over patient safety. There is a lack of methods to rationalize criteria design. In this study, we used data from a large research network to assess how adjustments of eligibility criteria can jointly affect generalizability and patient safety (i.e the number of serious adverse events [SAEs]). We first built a model to predict the number of SAEs. Then, leveraging an a priori generalizability assessment algorithm, we assessed the changes in the number of predicted SAEs and the generalizability score, simulating the process of dropping exclusion criteria and increasing the upper limit of continuous eligibility criteria. We argued that broadening of eligibility criteria should balance between potential increases of SAEs and generalizability using donepezil trials as a case study.

## Introduction

Clinical trials, especially randomized controlled trials (RCTs), are the current gold standard for measuring treatment effectiveness and safety,^1^ before a drug can be approved by the Food and Drug Administration (FDA). Trial sponsors and investigators often overemphasize the assessments of efficacy, and aim for good internal validity (i.e., how well the observed treatment effects are reflective of the true treatment effects in the study samples).^2^ On the other hand, the question of how well the study findings could be applied to the target patients in the real-world, referred to as external validity or generalizability, is often overlooked.^3^ Further, clinical trial designers often adopt eligibility criteria from existing similar studies, with no or little modifications, without sound scientific justifications. Many phase 3 trials continue to adopt the highly restrictive eligibility criteria used by their corresponding phase 1 and phase 2 trials^4^, resulting in study samples less representative of the real-world patient population who are in need of the treatments. For example, older adults are often excluded from, and hence underrepresented in cancer and Alzheimer’s Disease (AD) drug trials,^5,6^ despite being the primary target populations of these drugs. A recent study has found that among the most frequently prescribed drug classes with known differences in pharmacokinetics or contraindications for older adults, only 62% of the 113 initial approval documents had pharmacokinetic information for the elderly.^7^ Overly restrictive eligibility criteria will lead to low clinical trial generalizability, which will ultimately lead to low treatment effectiveness and increased risk of adverse events in certain population subgroups when the treatments are practiced in real-world patients. As a results, regulatory agencies such as the FDA had issued guidance on broadening eligibility criteria to increase the diversity of the clinical trial population during enrollment.^8^

In clinical trials, the study population includes patients who meet the eligibility criteria; the target population includes patients to whom the study findings will be applied; and the study sample population includes participants enrolled in the trials (***Figure 1***). Population representativeness measures the coverage of the study sample or study population over the target population with respect to study traits (e.g., demographic, diagnosis, and laboratory test results). Although population representativeness is a different concept from clinical trial generalizability, it is the key measure of a trial’s generalizability. To date, a number of methods and tools have been developed to quantify clinical trials’ population representativeness (or generalizability).^9^ These methods can be categorized into two major approaches: (1) sample-driven and often called *a posterior* generalizability, where these methods measure the representativeness the study samples (i.e., participants enrolled in clinical trials) over the target population, and (2) eligibility-driven and called *a prio*generalizability, where these methods measure the representativeness of the study population (i.e., patients who met the eligibility criteria) over the target population.^10^ Although the *a posterior* generalizability is important, it cannot be changed after the fact as the trial has already been concluded. In contrast, the *a priori* generalizability is driven by clinical trial’s eligibility criteria and can be tweaked when designing a trial. A clinical trial will have good *a priori* generalizability when its study population and target population share similar demographic and clinical characteristics. Among the available *a priori* generalizability assessment methods, the Generalizability Index of Study Traits (GIST) 2.0 is the best available quantitative, eligibility-driven generalizability measure. GIST 2.0 quantifies the population representativeness using eligibility criteria and data from the real-world target population.^11^ It measures the proportion of potentially eligible patients across multiple trial eligibility criteria, while considering the relative importance of individual traits.^12^ GIST 2.0 has two components: the single GIST (sGIST) when considering individual criteria and the multi-GIST (mGIST) when considering all the criteria of a trial as well as their weights in a trial as a set. The sGIST and mGIST range from 0 to 1, where a higher score indicates a greater generalizability. GIST 2.0 has been validated in previous studies, including our own.^13,14^

**Figure 1.**
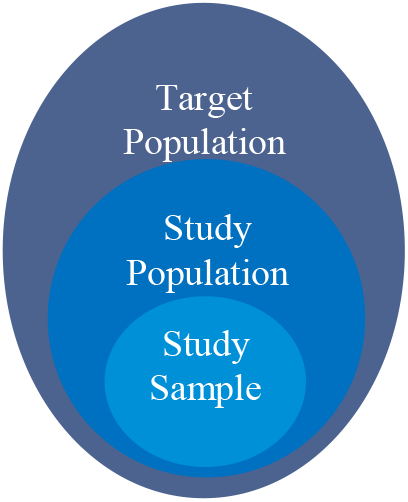
Population in clinical trials.

Although methods such as GIST are available for linking trial eligibility criteria and generalizability, it is unclear how broadening eligibility criteria will simultaneously impact trial generalizability and clinical outcomes in real-world patients. Investigators tend to use restrictive eligibility criteria for recruitment due to concerns over patient safety (e.g., fear of increased number of adverse events), but this is often done at the expense of trial generalizability with no clear data evidence. Therefore, it is crucial that we examine how *a priori* generalizability and the number of adverse events vary with adjustments to trial eligibility criteria so that a balance between internal and external validity can be identified. To our knowledge, no methods or tools are available to support and rationalize the eligibility criteria development process in clinical trial design through balancing generalizability and patient safety.

In this study, we aimed to analyze how broadening trial eligibility criteria will simultaneously impact trial generalizability, as measured by GIST, and clinical outcomes, as measured by serious adverse events (SAEs) using real-world data (RWD) from a large clinical data research network. We focused on Alzheimer’s disease (AD) patients who took the FDA-approved donepezil (Aricept), the most widely used drug for AD treatment. We obtained RWD data from the OneFlorida Clinical Research Consortium, a statewide clinical data repository that contains RWD, including electronic health records (EHRs) and administrative claims data, for over 14 million (> 50%) Floridians. We first built models to predict the number of donepezil-related SAEs using patients’ demographic and clinical characteristics. Then, we illustrated several scenarios in which we adjusted eligibility criteria and observed how the number of SAEs and trial generalizability changed at the same time. This study provided the initial evidence on how trial generalizability and clinical outcomes can be jointly affected by the adjustments of eligibility criteria and subsequently used as justification for broadening eligibility criteria as advocated by the FDA.

## Methods

### Data source and the overall patient cohort

The overall patient cohort for this study included patients who were diagnosed with AD and treated with donepezil. Donepezil is an acetylcholinesterase inhibitor under the brand name Aricept, which has known efficacy in patients with mild, moderate, and severe AD. We obtained individual-level patient data between January 2012 and March 2019 from the OneFlorida Clinical Research Consortium. OneFlorida is a statewide clinical research data repository that contains linked administrative claims and EHR data, including diagnoses, procedures, medications, and patient demographics, for approximately 14.4 million (> 50%) Floridians. OneFlorida was one of the 9 clinical data research network funded by the Patient-Centered Outcomes Research Institute (PCORI), contributing to the national Patient-centered Clinical Research Network (PCORnet). The OneFlorida data follow the PCORnet Common Data Model (CDM) that contains 22 data domains. We identified AD patients using International Classification of Disease, 9^th^ and 10^th^, Clinical Modification (ICD-9/10-CM) codes (i.e., ICD-9-CM: 331.0; ICD-10-CM: G30.0, G30.1, G30.8, and G30.9). Patients whose donepezil prescriptions were before their AD diagnoses were excluded from the study. Patients whose first donepezil prescription was within 90 days of their first encounter date in OneFlorida were also excluded to ensure a sufficient observation period. We identified 2,096 unique AD patients who were eligible for our study and extracted their data from OneFlorida. The selection of the overall patient cohort of our study is illustrated in **Figure 2**.

**Figure 2.**
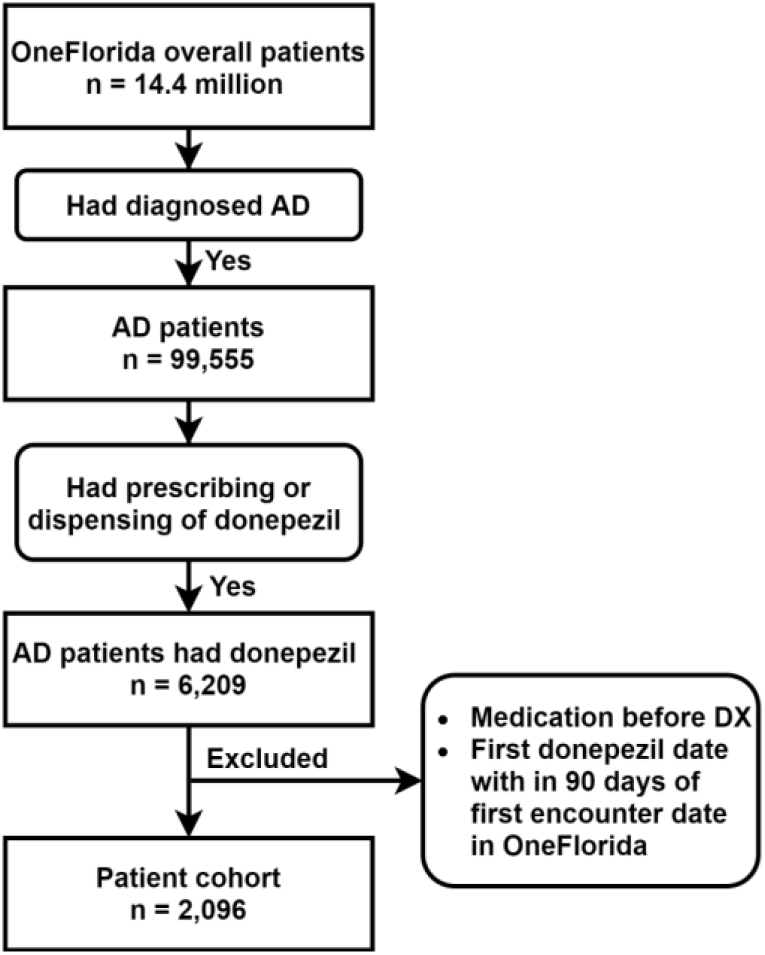
Selection of the overall patient cohort.

### Prediction model for the number of SAEs on AD patients treated with donepezil

To explore how adjustments to eligibility criteria affect the number of SAEs in the target population, we first built a prediction model for the number of SAEs on AD patients treated with donepezil, considering study traits (e.g., age) extracted from eligibility criteria as predictors. We also considered as model predictors other AD-related risk factors (e.g., chronic conditions) that can contribute to the number of SAEs. We argue that these additional predictors need to be considered as potential eligibility criteria in future trials.

#### Predictor variables

We first extracted all the eligibility criteria in all US-based Phase 3 donepezil AD trials on *ClinicalTrials*.*gov*. We then extracted study traits corresponding to each eligibility criterion from the OneFlorida EHR data as model predictors. For example, exclusion criterion “*patients with psychiatric disorders affecting the ability to assess cognition such as schizophrenia, bipolar or unipolar depression*” was converted to two binary study traits, (1) having schizophrenia and (2) having bipolar or unipolar depression, which were subsequently extracted from the EHR data using ICD codes for each patient. In addition, we used the chronic condition algorithms from the Centers for Medicare & Medicaid Services (CMS) Chronic Conditions Data Warehouse (CCW) to extract chronic conditions from the EHR data as model predictors.^15^ As shown in ***Figure 3***, we defined the observation window as the period before patients’ first donepezil prescription. Patient should have more than 90 days of data in the observation window (i.e., the patient shall have an encounter in the OneFlorida network 90 days before the first donepezil prescription). All predictor variables were extracted from the OneFlorida data within the observation window. To determine donepezil use, we extracted donepezil prescribing and dispensing data using RxNorm CUI codes and National Drug Codes (NDCs) and identified the first and last date of donepezil use for each patient.

**Figure 3.**
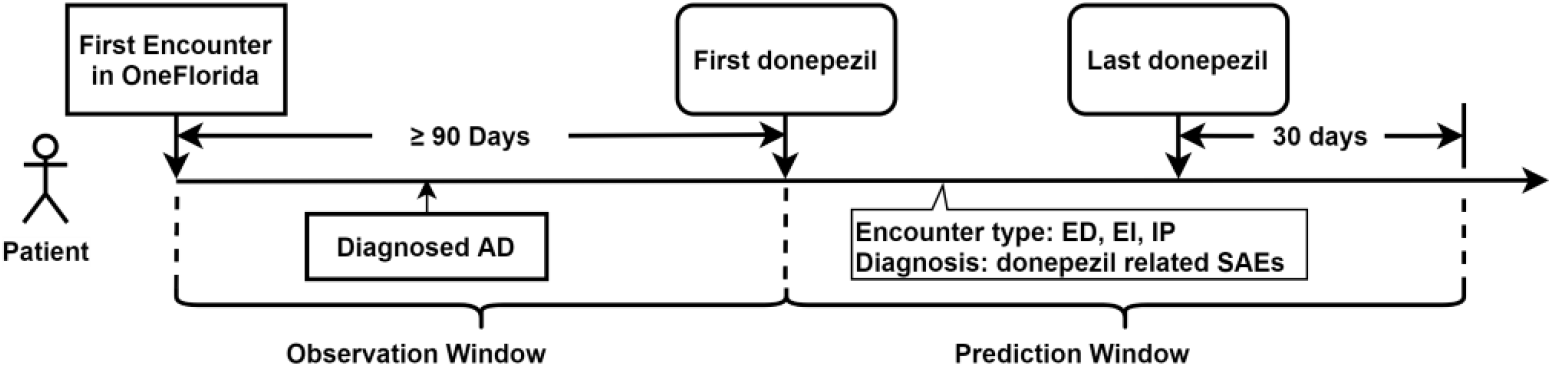
Definition of the observation window and prediction window.

#### Outcome variables

Our outcome variable was the number of SAEs occurred after donepezil use. To define SAEs, we first reviewed the drug label of donepezil (brand name Aricept) obtained from the DailyMed database and extracted the adverse events (AEs) from the warnings and adverse reactions sections. We also extracted and summarized AEs listed in all the completed donepezil-related AD trials that had results on *ClinicalTrial*.*gov*. We compiled a list of AEs from these two sources and identified 279 corresponding ICD-9-CM codes and 292 ICD-10-CM codes. Based on the AE severity grading scale defined in the Common Terminology Criteria for Adverse Events (CTCAE), AEs leading to hospitalization or prolongation of hospitalization are grade 3 AEs and were considered as SAEs in our study. Therefore, we identified encounters that had AE relevant ICD codes and subsequently had emergency department visits (ED), ED visits followed by inpatient hospital stays (EI), and inpatient hospital stays (IP) for each patient during the prediction window. The prediction window was defined as the period after the first donepezil prescription date but before the last donepezil prescription date plus 30 days (**Figure 3**). There were two limitations with our SAE definition: (1) we would miss certain SAEs, such as AEs that led to mortality because mortality data in OneFlorida was sparse, and (2) there was no explicit causal relationship between taking donepezil and the subsequent SAEs. Nevertheless, our SAE definition was reasonable because (1) the FDA AE guideline required reporting of all AEs after the treatment, not just those directly caused by the treatment, and (2) our analyses considered the average number of SAEs relatively across patient populations.

#### Prediction model

The number of SAEs is count data with only non-negative integer values and excessive zeros. Both the zero-inflated Poisson (ZIP) and zero-inflated negative binomial (ZINB) model can be used for this kind of outcomes. The ZINB model is a better choice when the outcome variable is overdispersed (i.e., the variance is much larger than the mean). Thus, we first examined the dispersion parameter of SAE counts to decide which of the two models was appropriate. Then, we built the model using all predictors defined above. We compared the model predicted probabilities to the true distributions to assess model fit. The prediction model was subsequently used as a basis for adjusting eligibility criteria while observing changes in predicted SAEs.

### Scenarios of eligibility criteria adjustments

To rationalize the adjustments of eligibility criteria, we explored how broadening eligibility criteria jointly impacted trial generalizability and SAE. To better illustrate the criteria broadening process, we used the pivot Phase 3 donepezil trial for AD, “*Comparison of 23 mg Donepezil Sustained Release (SR) to 10 mg Donepezil Immediate Release (IR) in Patients With Moderate to Severe Alzheimer’s Disease”* (NCT00478205) as a starting point for constructing a list of eligibility criteria for a hypothetical trial design. As AD trials often exclude patients with chronic conditions, we thus considered chronic conditions defined in the CMS CCW Chronic Condition algorithms as potential exclusion criteria, noting that some chronic conditions were already explicitly listed in NCT00478205 as exclusion criteria. To compute the trial’s generalizability scores (sGIST and mGIST), we defined the target population as the AD patients who were treated with donepezil in the OneFlorida data and used the original trial eligibility criteria to identify study population (i.e., those who met the criteria and eligible for the study in target population). The study population was defined as the eligible group and those who were not in study population but in the target population were defined as the non-eligible group (i.e., AD patients who were treated with donepezil but did not meet the trial eligibility criteria). For each criterion, a sGIST score could be calculated, with a lower sGIST score meaning the criterion was more stringent and thus excluded more patients from the study population compared to other criteria. An mGIST score could also be calculated for a trial, considering all eligibility criteria combined as well as the weights of the different study traits. A higher mGIST would mean the trial had a higher population representativeness, and thus better generalizability.

We considered two scenarios of eligibility criteria adjustments: (1) determine whether a binary (exclusion) criterion should be included or removed; and (2) determine the optimum range of a continuous criterion. To simplify the discussion, we did not used the terms inclusion and exclusion criteria; instead, we only considered the actual effects of the criteria – whether participants with certain study traits should be included in the trial or not.

In the first scenario, we considered how broadening binary criteria (i.e., disease diagnosis) impacted GIST and SAEs. We computed the sGIST score for each criterion and mGIST score for the hypothetical trial based on NCT00478205. To assess the overall effect of a criterion-corresponding study trait on SAEs, we used the prediction model to compute the predicted mean number of SAEs for each study trait. If the sGIST score of the criterion was small and the corresponding study trait had a weak association with number of SAEs (i.e., the criterion was limiting trial generalizability but had little effect on the number of SAEs), the criterion could potentially be eliminated. Further, we removed individual study traits from the original trial eligibility criteria one at a time and assessed its impact on the mGIST and predicted mean number of SAEs of the study population for the eligible and non-eligible groups, respectively. Through monitoring the joint changes of mean SAEs and mGIST scores, we can observe if removing certain criteria is worthwhile considering the tradeoff between the mean number of SAEs in the target population (both eligible and non-eligible groups) and the trial generalizability in terms of mGIST score.

In the second scenario, we considered how broadening a continuous criterion (i.e., age) jointly impacted the mGIST score and predicted mean number of SAEs. In the trial NCT00478205, the age criterion was set to be between 45 and 90 years old. We broadened the age criterion by sequentially increasing the upper age limit, one year at a time, to 100 years. At each iteration of age increase, we computed the trial’s mGIST score and predicted mean number of SAEs. The mGIST scores would increase as the range of the age criterion enlarges; meanwhile, as it enlarges, if the model adjusted mean number of SAEs of patients within the age criterion (holding other criteria unchanged) is not significantly higher, the upper limit of the age criterion could be enlarged.

## Results

### Patient cohort characteristics

We identified 2,096 unique patients who were eligible for our study. Among these patients, 1,351 (64.5%) had zero SAEs and 745 (35.5%) had at least one SAE. The overall mean age at AD diagnosis was 77.2, and the overall mean age at first donepezil medication was 78.2. Patients who had any SAEs were slightly older than those who had no SAEs (77.7 vs. 77.0; *p* = 0.0973). There were more female than male patients (63.4% vs. 36.6%; *p* = 0.4499). Over 50% of the patients were non-Hispanic white. The percentage of the non-Hispanic black patients was higher in the patient group having SAEs compared to the no SAE group. The characteristics of the study population is summarized in **Table 1**.

**Table 1.** Patient characteristics of the target population in OneFlorida.

|  | Overall<br>(N=2,096) |  | # of SAEs = 0<br>(N=1,351) |  | # of SAE > 0<br>(N=745) |  | P value* |
| --- | --- | --- | --- | --- | --- | --- | --- |
|  | N (or<br>Mean) | % (or SD) | N (or<br>Mean) | % (or SD) | N (or<br>Mean) | % (or SD) |  |
| Age at AD diagnosis | 77.2 | 9.7 | 77.0 | 9.5 | 77.7 | 9.9 | 0.0973<sup>a</sup> |
| Age at first donepezil | 78.2 | 9.7 | 78.0 | 9.6 | 78.5 | 9.9 | 0.2303<sup>a</sup> |
| Gender |  |  |  |  |  |  | 0.4499<sup>b</sup> |
| Female | 1328 | 63.4% | 848 | 62.8% | 480 | 64.4% |  |
| Male | 768 | 36.6% | 503 | 37.2% | 265 | 35.6% |  |
| Race/Ethnicity |  |  |  |  |  |  | <.0001<sup>b</sup> |
| NHW | 1050 | 50.1% | 706 | 52.3% | 344 | 46.2% |  |
| NHB | 408 | 19.5% | 215 | 15.9% | 193 | 25.9% |  |
| Hispanic | 563 | 26.9% | 376 | 27.8% | 187 | 25.1% |  |
| Other | 75 | 3.6% | 54 | 4.0% | 21 | 2.8% |  |
|  | **Median** | **IQR** | **Median** | **IQR** | **Median** | **IQR** |  |
| Number of donepezil prescriptions | 2.0 | (1.0, 3.0) | 1.0 | (1.0, 2.0) | 2.0 | (1.0, 4.0) | <.0001<sup>c</sup> |
| Number of months on donepezil | 0.0 | (0.0, 6.6) | 0.0 | (0.0, 0.9) | 1.6 | (0.0, 17.0) | <.0001<sup>c</sup> |
| *The p value was for the comparison between the # of SAEs = 0 and # of SAE > 0 group |  |  |  |  |  |  |  |
| <sup>a</sup>two samples T test; |  |  |  |  |  |  |  |
| <sup>b</sup>Chi-square test; |  |  |  |  |  |  |  |
| <sup>c</sup>Wilcoxon rank sum test; |  |  |  |  |  |  |  |

### Analysis of donepezil AD trial eligibility criteria

We identified a total of 5 Phase 3 trials conducted in the U.S. testing donepezil for treating AD (NCT00478205, NCT00566501, NCT00428389, NCT00096473, and NCT00000173) and extracted 113 eligibility criteria (54 inclusion and 52 exclusion criteria). On average, each donepezil AD trial had 23 criteria. Some criteria could be decomposed into multiple sub-criteria (e.g., “*hypertension and cardiac disease must be well-controlled*” could be decomposed into “*well-controlled hypertension*” and “*well-controlled cardiac disease*”). We decomposed these eligibility criteria and extracted the core elements of each criterion. Many of the eligibility criteria were fundamentally similar (e.g., “*age > 40*” and “*age > 45*” both discussed a core element on patient age). We considered the smallest core elements of criteria as individual study traits and extracted 193 unique study traits out of the 113 eligibility criteria. However, not all study traits were computable against our OneFlorida patient database. For example, there were 2 inclusion and 5 exclusion criteria related to the availability of caregivers to the patients (e.g., “*caregiver must have regular contact with the patient*”), which were not computable using OneFlorida data. We found that 60 (31.1%) of the study traits were not computable. The main reasons were: (1) the trait was based on subjective information (e.g., requiring “*informed consent*”, or health conditions that require investigator’s judgement); and (2) the data elements were not available in the OneFlorida data (e.g., information on whether a patient “*lives in assisted living facility*” was not available in the structured OneFlorida data).

### Prediction model for the number of SAEs

The dispersion parameter (0.885; 95% confidence interval [CI]: 0.730, 1.071) for the SAEs was statistically significant from zero, indicating that ZINB regression, rather than ZIP regression, should be used for modeling. As shown in ***Figure 4***, the ZINB model predicted probabilities being close to the observed relative frequencies indicated a good fit.

**Figure 4.**
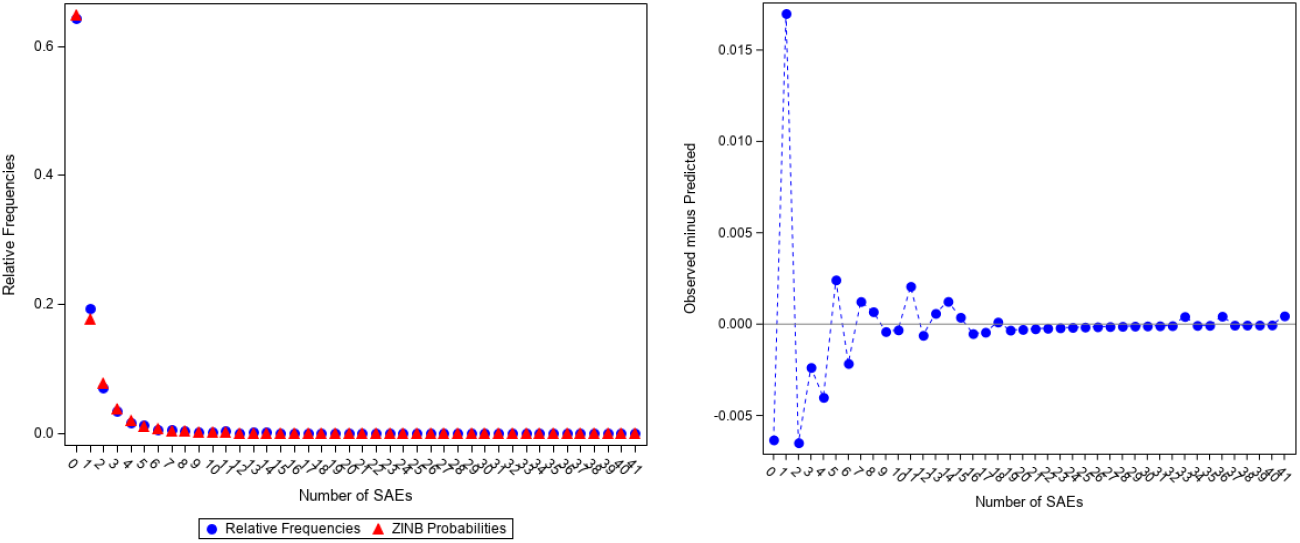
ZINB Predicted Probabilities and Observed Relative Frequencies.

The ZINB model estimates were shown in **Table 2**.

**Table 2.** Zero-inflated negative binomial model for predicting number of SAEs.

| **Part 1: logistic part for excessive zero (i.e., having no SAE)** |  |  |  |
| --- | --- | --- | --- |
| **Parameter** | **Odds Ratio (OR)** | **95% Confidence Interval** | **p-value** |
| Age at first donepezil | 0.972 | (0.929, 1.017) | 0.2181 |
| Male vs Female | 1.739 | (0.711, 4.255) | 0.2256 |
| Race/Ethnicity |  |  |  |
| Hispanic vs NHW | 0.204 | (0.079, 0.531) | 0.0011 |
| NHB vs NHW | 0.659 | (0.232, 1.867) | 0.4321 |
| Number of donepezil prescription | 0.027 | (0.003, 0.231) | 0.0010 |
| Number of months on donepezil | 0.966 | (0.834, 1.119) | 0.6448 |
| Chronic conditions * (Only $p < 0.05$ are shown here) | | | |
| Anemia | 0.268 | (0.097, 0.74) | 0.0110 |
| Ischemic Heart Disease | 0.122 | (0.029, 0.52) | 0.0044 |
| **Part 2: negative binomial part** |  |  |  |
| **Parameter** | **Estimate** | **95% Confidence Interval** | **p-value** |
| Age at first donepezil | 0.998 | (0.988, 1.007) | 0.5995 |
| Male vs Female | 1.072 | (0.867, 1.326) | 0.5213 |
| Race/Ethnicity |  |  |  |
| Hispanic vs NHW | 0.874 | (0.699, 1.093) | 0.2369 |
| NHB vs NHW | 1.285 | (1.042, 1.584) | 0.0190 |
| Number of donepezil prescription | 1.104 | (1.071, 1.139) | <.0001 |
| Number of months on donepezil | 1.023 | (1.017, 1.03) | <.0001 |
| Chronic conditions * (Only $p < 0.05$ are shown here) | | | |
| Chronic obstructive pulmonary disease (COPD) | 1.397 | (1.134, 1.719) | 0.0016 |
| Hyperlipidemia | 0.756 | (0.624, 0.916) | 0.0042 |
| Hypertension | 1.517 | (1.202, 1.914) | 0.0004 |
| Anxiety disorder | 1.368 | (1.115, 1.679) | 0.0027 |

The first part of the ZINB model was a logistic regression model for estimating the probability of having no SAE. Age and gender were not statistically significant in this part of the model. Hispanics had a lower probability of having no SAE compared to non-Hispanic whites (OR = 0.204; *p* = 0.0011). The number of donepezil prescriptions was a significant predictor (OR = 0.027; *p* = 0.0010), indicating that having more donepezil doses was associated with a lower probability of having no SAE. In terms of chronic conditions, patients with anemia and ischemic heart disease had significant lower odds of having no SAE (OR = 0.268; *p* = 0.0110 and OR = 0.122; *p* = 0.0044).

The second part of the ZINB model was a negative binomial regression, estimating the expected number of SAEs conditioned on having at least one SAE. Age at first donepezil prescription, gender, and race/ethnicity were not statistically significant in this part of the model. The number of donepezil prescriptions and the number of months on donepezil were significant predictors, indicating that increasing 1 donepezil prescription would increase the number of SAEs by 1.104 – 1 = 0.104 (beta =, *p* < 0.0001) and increasing 1 month of being on donepezil would increase the number of SAEs by 1.023 – 1 = 0.023 (beta =; *p* < 0.0001). In terms of chronic conditions, chronic obstructive pulmonary disease (COPD) had an estimate of 1.397 (*p* = 0.0016), indicating having COPD would increase the number of SAEs. Hyperlipidemia had an estimate smaller than 1, indicating patients with hyperlipidemia would have 1 – 0.756 = 0.244 (*p* = 0.0042) fewer SAEs. Patients with hypertension would have 1.517 – 1 = 0.517 (*p* = 0.0004) more SAEs than those without hypertension. Anxiety disorder had an estimate of 1.368 (*p* = 0.0027), indicating patients with anxiety disorder would also have 0.368 more SAEs.

### The relationships among eligibility criteria, SAEs, and trial generalizability

The original donepezil trial NCT00478205 had 40 inclusion and exclusion criteria, with 4 criteria about caregivers. Based on the eligibility criteria from NCT00478205 and the 27 chronic conditions in the CMS CCW algorithms, we constructed a hypothetical trial design with 18 criteria as shown in **Table 3**. Excluding non-computable criteria, we extracted 31 study traits from the 18 criteria. Note that for simplification, we did not use individual study traits in this part of the analysis. For example, strictly speaking, “*visual or hearing impairment*” were two different study traits: “*visual impairment*” and “*hearing impairment*”; nevertheless, we combined these two as one exclusion criteria.

**Table 3.** Eligibility criteria of a hypothetical trial constructed based on NCT00478205.

| **Num** | **Short Name** | **Inclusion/**<br>**Exclusion** | **Criteria Definitions** |
| --- | --- | --- | --- |
| 1 | Age | Inclusion | age at first donepezil date 45 - 90 |
| 2 | Donepezil days | Inclusion | days on donepezil ≥ 90 days |
| 3 | Visual/hearing Impairment | Exclusion | patients with visual impairment or hearing impairment* |
| 4 | Cardiac diseases | Exclusion | patients with acute myocardial infraction, atrial fibrillation, heart failure, or ischemic heart disease* |
| 5 | Uncontrolled Hypertension | Exclusion | patients with hypertension and have systolic blood pressure $> 140$ or diastolic blood pressure $> 90$ in recent 3 months* |
| 6 | Uncontrolled diabetes | Exclusion | patients with diabetes and have HbA1c $> 7\%$ in recent 3 months* |
| 7 | Other AD treatments | Exclusion | patients taken any of Tacrine, Pyridostigmine, Galantamine, Isoflurophate, Demecarium, Physostigmine, Rivastigmine, Edrophonium, or Ambenonium |
| 8 | Dementias other than AD | Exclusion | patients had no diagnosis of AD but had diagnosis of other dementias |
| 9 | Parkinson's disease | Exclusion | patients diagnosed with Parkinson's disease |
| 10 | Schizophrenia | Exclusion | patients with schizophrenia* |
| 11 | Depression | Exclusion | patients with depression* |
| 12 | Sleep disorder | Exclusion | patients diagnosed with sleep disorder |
| 13 | Drug use disorders | Exclusion | patients with drug use disorders* |
| 14 | Alcohol use disorders | Exclusion | patients with alcohol use disorders* |
| 15 | Conditions affect absorption, distribution, or metabolism of the study medication | Exclusion | patients diagnosed with any of inflammatory bowel disease, gastric or duodenal ulcers, or hepatic disease |
| 16 | Cancer | Exclusion | patients with a history of cancer (does not include basal or squamous cell carcinoma of the skin, benign prostatic hyperplasia) within 5 years* |
| 17 | Use antidepressants | Exclusion | patients prescribed with any of amitriptyline, clomipramine, doxepin, imipramine, trimipramine, protriptyline, amoxapine, desipramine, or nortriptyline |
| 18 | Fecal/urinary incontinence | Exclusion | patients diagnosed with fecal or urinary incontinence |

\*These criteria were constructed based on CMS CCW chronic condition algorithms.

We summarized the predicted mean number of SAEs and sGIST scores for each of the 16 exclusion criteria in **Figure 5**. Patients with Parkinson’s disease had the lowest mean number of SAEs at 0.65. Patients who had taken antidepressant medications had the highest mean number of SAEs at 1.87. However, only 12 patients in the OneFlorida data had taken antidepressants. Patients who had alcohol use disorder also had a high mean number of SAEs at 1.70. In terms of sGIST scores, the exclusion criterion of cardiac diseases had the lowest score of 0.578, indicating it was the most stringent criterion. Exclusion based on depression and uncontrolled hypertension also had low sGIST scores of 0.716 and 0.768, respectively. Exclusion based on the use of antidepressants had the highest sGIST score of 0.991. Exclusion based on uncontrolled diabetes had a sGIST score of 0.964. For alcohol use disorder, the sGIST was 0.962.

**Figure 5.**
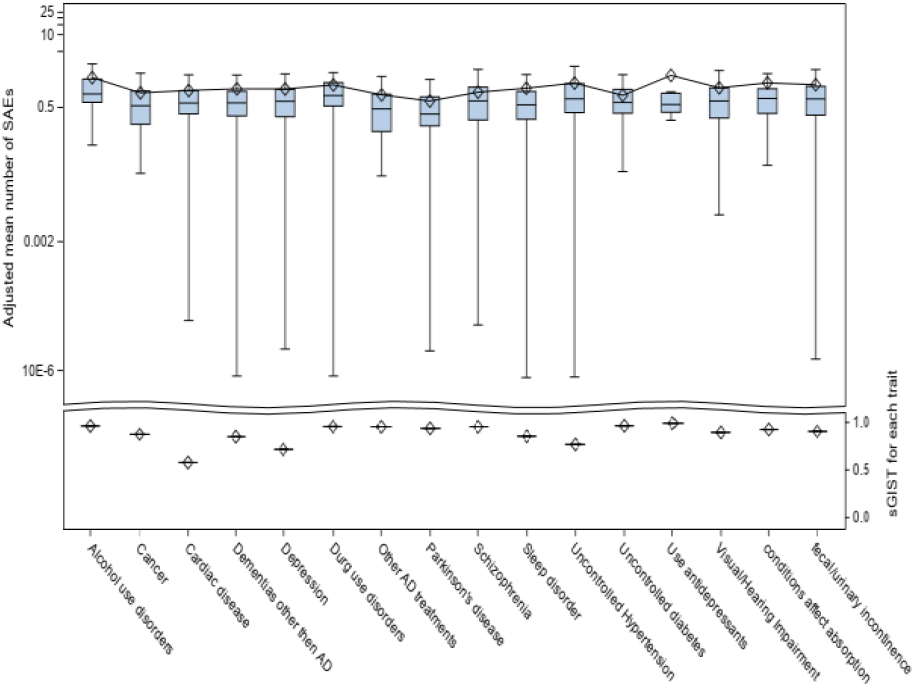
Model adjusted number of SAEs (upper) and sGIST (bottom) for each of the 16 exclusion criteria.

The effects of removing an individual exclusion criterion on the number of SAEs and the mGIST score were shown in **Table 4**. Out of the 2,096 AD patients treated with donepezil, 373 met the eligibility criteria of the original hypothetical study (i.e., the eligible group) with a predicted mean number of SAEs of 0.66. The non-eligible group had a higher predicted mean number of SAEs of 0.99. The mGIST score for the original hypothetical trial was 0.062. As an example, if we broadened the eligibility criteria by removing the exclusion criterion of cardiac diseases that had the lowest sGIST of 0.578, the mGIST score of the trial would increase to 0.078, while the number of patients in the eligible group increased from 373 to 503 and the adjusted mean number of SAEs for eligible and non-eligible groups would increase from 0.66 to 0.67 and from 0.99 to 1.01, respectively. On the other hand, dropping the criterion of uncontrolled hypertension would lead to a smaller mGIST gain (i.e., from 0.062 to 0.074) but significantly increase the mean number of SAEs in the eligible group from 0.66 to 0.83. Based on our results on SAE and mGIST, one can rationalize the choice of dropping cardiac diseases versus dropping uncontrolled hypertension as exclusion criteria.

**Table 4.** Population size, model adjusted mean number of SAEs, and mGIST when dropping individual exclusion criteria, independently, stratified by eligible vs. non-Eligible group.

| Independently dropping individual exclusion criteria | Eligible |  | non-Eligible |  | sGIST* | mGIST |
| --- | --- | --- | --- | --- | --- | --- |
|  | Population Size (N) | # of Mean SAEs | Population Size (N) | # of Mean SAEs |  |  |
| 00.Original | 373 | 0.66 | 1642 | 0.99 | . | 0.062 |
| **The lowest sGIST scores after dropping these exclusion criteria** |  |  |  |  |  |  |
| 01.Drop Cardiac disease | 503 | 0.67 | 1512 | 1.01 | 0.578 | 0.078 |
| 02.Drop Depression | 431 | 0.68 | 1584 | 0.99 | 0.716 | 0.074 |
| 03.Drop Uncontrolled Hypertension | 431 | 0.83 | 1584 | 0.95 | 0.768 | 0.074 |
| **The highest sGIST scores after dropping these exclusion criteria** |  |  |  |  |  |  |
| 13.Drop Drug use disorders | 378 | 0.66 | 1637 | 0.99 | 0.955 | 0.062 |
| 14.Drop Alcohol use disorders | 378 | 0.66 | 1637 | 0.99 | 0.962 | 0.063 |
| 15.Drop Uncontrolled diabetes | 378 | 0.66 | 1637 | 0.99 | 0.964 | 0.062 |
| 16.Drop Use antidepressants | 373 | 0.66 | 1642 | 0.99 | 0.991 | 0.062 |
| *sGIST score of the specific exclusion criterion before dropping the exclusion criterion. |  |  |  |  |  |  |

**Table 5** shows a different scenario of dropping exclusion criteria, where we assessed the impact of dropping multiple exclusion criteria on the mean number of SAEs and mGIST score of the trial. It was clear as we dropped additional exclusion criteria, both the population size of the eligible group and the mGIST score of the trial increased. However, the predicted mean number of SAEs also increased, highlighting the need to find a balance between gaining trial generalizability and potentially increasing SAEs.

**Table 5.** Population size, model adjusted mean number of SAEs, and mGIST when dropping combined individual exclusion criteria, sequentially, stratified by eligible vs. non-Eligible group.

| Subsequently dropping criterion | Eligible |  | non-Eligible |  | mGIST |
| --- | --- | --- | --- | --- | --- |
|  | Population Size (N) | # of Mean SAEs | Population Size (N) | # of Mean SAEs |  |
| 00.Original | 373 | 0.66 | 1642 | 0.99 | 0.062 |
| 01.Drop Cardiac disease | 503 | 0.67 | 1512 | 1.01 | 0.078 |
| 02.Drop Depression | 603 | 0.70 | 1412 | 1.02 | 0.096 |
| 03.Drop Uncontrolled Hypertension | 744 | 0.83 | 1271 | 0.98 | 0.120 |
| 04.Drop Dementias other than AD | 865 | 0.82 | 1150 | 1.00 | 0.141 |
| 05.Drop Sleep disorder | 972 | 0.85 | 1043 | 1.00 | 0.157 |
| 06.Drop Cancer | 1101 | 0.83 | 914 | 1.04 | 0.175 |
| 07.Drop Visual/Hearing Impairment | 1206 | 0.85 | 809 | 1.04 | 0.194 |
| 08.Drop fecal/urinary incontinence | 1320 | 0.90 | 695 | 0.97 | 0.214 |
| 09.Drop Conditions affect absorption | 1413 | 0.93 | 602 | 0.92 | 0.232 |
| 10.Drop Parkinson's disease | 1499 | 0.91 | 516 | 0.97 | 0.241 |
| 11.Drop Other AD treatments | 1575 | 0.91 | 440 | 0.99 | 0.256 |
| 12.Drop Schizophrenia | 1645 | 0.90 | 370 | 1.04 | 0.260 |
| 13.Drop Drug use disorders | 1703 | 0.91 | 312 | 1.01 | 0.269 |
| 14.Drop Alcohol use disorders | 1778 | 0.94 | 237 | 0.80 | 0.280 |
| 15.Drop Uncontrolled diabetes | 1846 | 0.94 | 169 | 0.78 | 0.290 |
| 16.Drop Use antidepressants | 1856 | 0.95 | 159 | 0.69 | 0.292 |

**Figure 6** illustrates the second scenario of eligibility criteria adjustments, where we aimed to find the optimal range of a continuous criterion. Using the age criterion (i.e., patients from 45 years old to 90 years old) as an example, we gradually increased the upper limit of the age criterion from 40 to 100. Considering patients’ age at first donepezil prescription, the predicted mean number of SAEs for each unit increase of the upper age limit, the sGIST score of the age criterion, and mGIST score of the trial were plotted in **Figure 6**. As the upper limit of the age criterion increased, the number of SAEs slightly increased at the beginning and then decreased slightly, but essentially vibrated between 1.3 to 1.5. The confidence interval of the model predicted mean number of SAEs were large between 50 and 60 as well as between 90 and 100, but relatively narrow between 70 and 80, because we had more data of patients between 70 and 80. Both the sGIST score of the age criterion and the overall mGIST of the trial increased, at first, quickly from 70 to 80 and then slowed down after around 85. Considering both the GIST and the predicted mean number of SAEs, it may be beneficial to increase the upper limit of the age criterion to 100 because the increase in trial generalizability was not accompanied with significant increase in mean number of SAEs.

**Figure 6.**
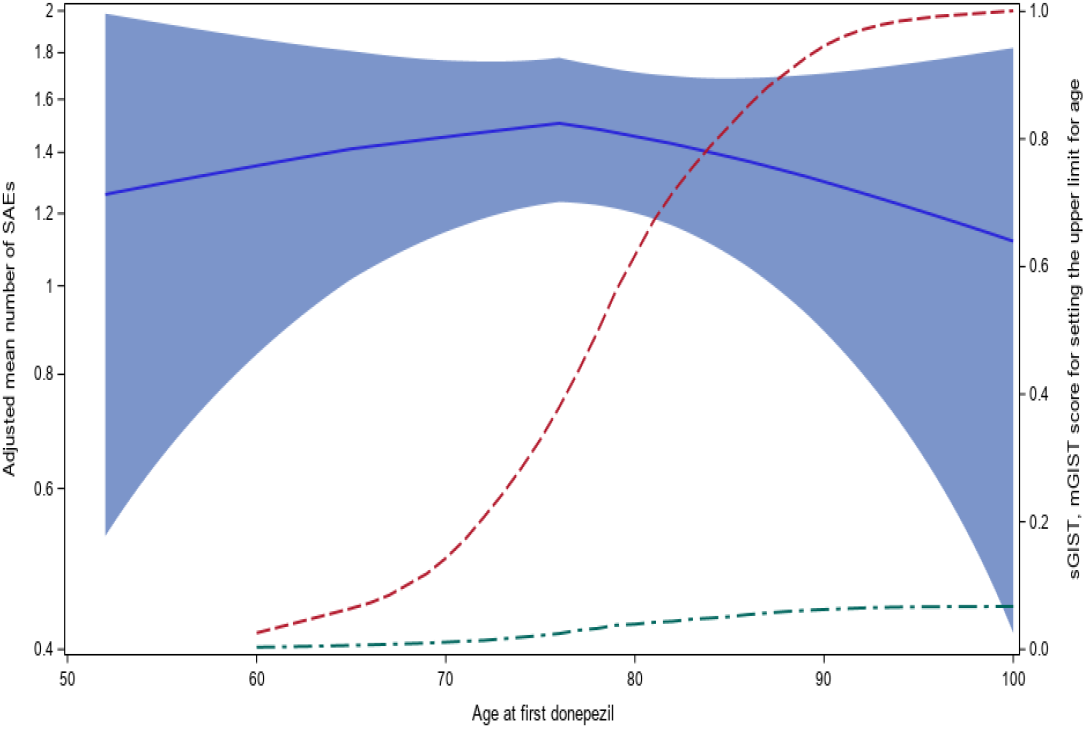
Model adjusted mean number of SAEs across age (blue solid line, the band were 95% confidence interval) sGIST score for age criterion with lower limit of 40 (red dash line), and mGIST score of the trial (green dash line).

## Discussion and conclusion

Our study demonstrated that adjusting clinical trial eligibility criteria would simultaneously impact trial generalizability and SAEs in the target population using RWD from a large clinical data network—OneFlorida. We also demonstrated the potential decision processes of rationalizing both categorical and continuous eligibility criteria with RWD. By examining the predicted number of SAEs for subgroup defined by each criterion, the subgroup with a lower risk of having SAE should be allowed to participate. By examining the sGIST scores, the eligibility criterion has the most stringency could potentially be dropped. Nevertheless, adjustments to eligibility criteria should consider both the generalizability of the trial (reflected by the GIST scores) and the predicted mean number of SAEs simultaneously. For categorical traits like chronic conditions, usually used as exclusion criteria, if dropping such a trait (i.e., so that patients with that certain disease would be included in the trial) would largely increase the number of SAEs but gain little in trial generalizability, it may not be a good idea to do so. For continuous trait like age, we shall broaden the age limits to include as many patients as possible, especially older adults, but without increasing the risk of potential SAEs. Studies had shown that older patients, especially for those above 80, were under-represented in existing AD trials.^6^ As demonstrated in our study, for the donepezil trial for treating AD patients, the patients who aged above 80 had a similar expected number of SAEs comparing to those who were younger; thus, increasing the upper age limit to include older participants should be allowed. In sum, eligibility criteria design of a trial should find the balance between manageable risks of adverse events for those eligible for the trial and the maximum trial generalizability.

Our approach of using RWD to rationalizing clinical trial eligibility criteria by linking them with a generalizability score and the number of SAEs can be easily applied to other clinical data networks that contain large collections of RWD. For other diseases and treatments, the same steps could be used to examine how adjustments to eligibility criteria can jointly impact trial generalizability and drug-related SAEs, which informs trial design. Clinical trials are typically conducted in phases, where one could use our approach and data collected from early phase trials (e.g., phase 1 and 2 trials) to inform the design, especially eligibility criteria design, of later phase trials (e.g., phase 3 trials). Such an approach will yield a high return on investments, where phase 3 trials can be tailored to have the greatest generalizability with manageable participant risks.

Moreover, our study also shows the feasibility of using RWD to build a trial participant identification and recruitment tool. This tool would allow exploration of the potential target population and their characteristics, designing and tailoring the trial eligibility criteria, assessing the sample size of the study population, estimating the clinical outcomes (e.g., number of SAEs), and assessing the trial’s generalizability. With such a tool, RWD could be used to support trial design to narrow the population representativeness gap between the trial participants and real-world target patients. Additionally, such tool would also help assess participant risks in terms of SAEs when planning the trial.

Our study is not without limitations. First, we had no information on medication adherence because of the limitation of the EHR data, where only prescription data are available. We simply assumed that patients who were prescribed with the medication did take the medication. In future studies, being able to link EHR data with medication dispensing data can potentially alleviate this limitation. Second, a number of the eligibility criteria were not computable because of the limited availability of the data elements in structured OneFlorida data (e.g., MMSE scores). However, these data elements are often documented in free-text clinical narratives. In future studies, we can explore advanced natural language process (NLP) methods to extract these important data elements from unstructured clinical narratives. Moreover, in addition to SAEs, other clinical outcomes such as survival and treatment efficacy could be explored to enhance the decision processes.

In sum, tools and methods to support the design of eligibility criteria are in great need. Our ultimate goal is to build an easy-to-use eligibility criteria design tool that could rationalize the eligibility criteria by balancing clinical outcomes and trial generalizability with real-world data.

## Data Availability

The data is from the OneFlorida Clinical Research Consortium.

## Acknowledgements

This work was supported in part by NIH grants R21AG068717, R21AG061431, R01CA246418, UL1TR001427, and PCORI ME-2018C3-14754 and the OneFlorida Clinical Research Consortium (PCORI CDRN-1501-26692). The content is solely the responsibility of the authors and does not necessarily represent the official views of the NIH or PCORI.

